# H-MpoxNet: A Hybrid Deep Learning Framework for Mpox Detection from Image Data

**DOI:** 10.1101/2024.11.26.24318006

**Authors:** Sajal Chakroborty

**Affiliations:** Worcester Polytechnic Institute, Worcester, Massachusetts, USA

**Keywords:** Deep learning, Machine learning classifiers, Feature extraction, Mpox detection, Infectious disease, Image data analysis

## Abstract

Infectious diseases pose significant global threats to public health and economic stability by causing pandemics. Early detection of infectious diseases is crucial to prevent global outbreaks. Mpox, a contagious viral disease first detected in humans in 1970, has experienced multiple epidemics in recent decades, emphasizing the development of tools for its early detection. In this paper, we propose a hybrid deep learning framework for Mpox detection. This framework allows us to construct hybrid deep learning models combining deep learning architectures as a feature extraction tool with machine learning classifiers and perform a comprehensive analysis of Mpox detection from image data. Our best-performing model consists of MobileNetV2 with LightGBM classifier, which achieves an accuracy of 91.49%, precision of 86.96%, weighted precision of 91.87%, recall of 95.24%, weighted recall of 91.49%, F1 score of 90.91%, weighted F1-score of 91.51% and Matthews Correlation Coefficient score of 0.83.

## 1 Introduction

Mpox, also known as monkeypox (MPX), is caused by the Mpox virus (MPXV). It is a zoonotic disease, more specifically an infectious skin disease that can spread between animals and humans. It is a 200-250 nm brick-like or ovoid shaped double helix DNA virus that belongs to the Orthopoxvirus genus, Proxviriade family, and Chordopoxvirinae subfamily [68]. MPXV can be transmitted into the human body in several ways, from animal to human and human to human [68]. Moreover, it can be transmitted by contact with contaminated objects, a patient’s droplets, body fluids, and lesions [68]. After exposure to MPXV, it takes 3-17 days to develop symptoms, which can last nearly 2-5 weeks [15]. According to the Centers for Disease Control and Prevention, a few symptoms of Mpox are fever, swollen lymph nodes, headache, muscle aches and backache.

The first human case caused by MPXV was reported in 1970 [27, 43]. In 2022-23, there was a global outbreak of Mpox [31]. Before that, between 1970 and 2017, several outbreaks occurred in central and west Africa [53, 13, 24]. Clade I, previously known as the Congo Basin Clade or Central African Clade, and Clade II, previously known as West African Clade, were responsible for the outbreaks in Central and West Africa [14]. The first outbreak outside Africa occurred in the USA in 2003 [46, 14]. In 2023, 809 positive cases were reported in different states of the USA, and 1,945 have been reported up to September 09, 2024 [16]. In Figure 1, we present a scenario based on data collected from the websites of the “Centers for Disease Control and Prevention” and “Our World in Data” for different states in the USA and worldwide. From these reported cases, it is visible that developing tools for the early detection of Mpox has evolved into a significant research problem.

**Figure 1:**
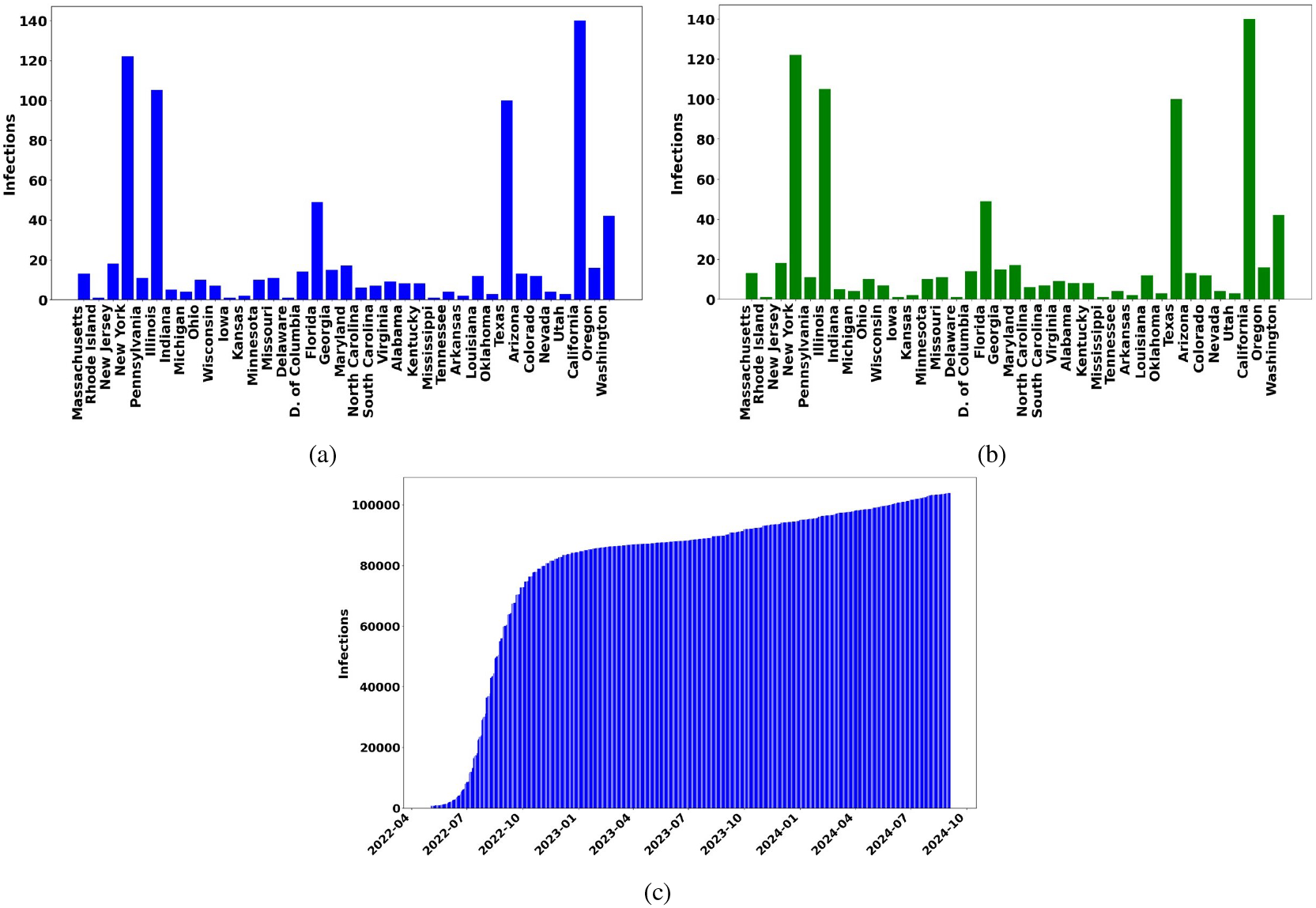
Mpox reported cases in the USA and Worldwide. Figures 1a and 1b illustrate the number of infected cases in 2023 and 2024, respectively, in different states of the USA, and 1c shows cumulative infected cases reported globally up to September 2024.

A widely used diagnostic tool for infectious diseases is Polymerase Chain Reaction (PCR) test [25]. It provides precise genetic analysis. However, PCR has risks of contamination, and limitations in detecting large or unknown sequences [69]. Moreover, it requires specialized equipment and expertise, making it less accessible in some resource-limited settings [69].

In recent years, deep learning (DL) techniques have become an alternative to detect infectious diseases. For a detailed review, one can see [11], [32], and [60]. While PCR is more likely inclined toward the lab resources and has a risk of contamination, DL techniques are mainly focused on biomedical images. DL techniques are data driven and a subclass of artificial intelligence [30]. It can predict patterns from large and complex datasets [30]. It involves training neural networks that can mimic the neural orientation of the human brain [30]. Most importantly, once the model is trained, it can be deployed on a simpler system to predict, classify, and generate output [30].

In addition to DL, there are machine learning (ML) tools for classification. For example, Support vector machine (SVM), Logistic regression, Random Forest (RF), Naïve bayes (NB). Due to computational efficiency these classifiers have found wide ranging applications in various fields [49, 56].

A common thread among these ML and DL techniques for disease detection is their use of biomedical images, such as histopathological ([45, 2, 4]) and MRI images ([7, 8, 40]). For instance, chest X-ray images have been used for early COVID-19 detection (see [59], [35], and [34]). Notably, these techniques do not pose any contamination risk, unlike PCR.

Although DL and ML techniques have demonstrated satisfactory performance in disease prediction, they require a large volume of images for model training [39]. Moreover, the training process demands high-performance computational resources [39]. Feature extraction from images is an initial and essential step for training and testing the models, which also requires significant computational time [39]. Therefore, developing a computational strategy to reduce this time has been an active area of research for a long period.

Furthermore, datasets could be imbalanced, and due to this, model prediction accuracy could be erroneous [18]. Consequently, developing a robust model could be very challenging. Most studies in literature have addressed this issue through data augmentation [61]. However, data augmentation for images is much more complex compared to tabular data. Some widely used techniques for image data augmentation include generative adversarial networks (GANs) and diffusion generative models [61]. However, the training process for these models is highly computationally expensive [61].

Various DL, ML, and hybrid methods have been proposed in prior studies to identify Mpox from skin lesion images. A few research gaps we have identified in these studies include: to develop robust models, most of these papers implement various data augmentation techniques; however, they did not address whether this augmentation introduces any bias. Since imbalanced data can produce erroneous accuracy, some other metrics that are widely used for imbalanced data were not explored. Finally, very few studies have shown a comprehensive analysis of various DL or ML models for Mpox detection. We elaborate on these research gaps in the literature review section.

The primary contributions of this paper to Mpox detection include:

i. We develop a hybrid DL framework, which combines a pretrained DL architecture with an ML classifier. We use pretrained DL architectures to extract features from images. After that, we train the ML model for Mpox prediction.
ii. We perform image augmentation using standard techniques that have been used in prior studies to develop robust hybrid models. To validate that these augmented images do not introduce any biases, we perform the Kullback-Leibler divergence test. To our knowledge, no prior studies have done this.
iii. We assess the robustness of our hybrid models through a comprehensive analysis, computing various metrics, including accuracy, precision, weighted precision, recall, weighted recall, F1 score, weighted F1 score, receiver operating characteristic (ROC) curve, and area under the curve (AUC). Since imbalanced data can impact accuracy, we also compute the Matthews correlation coefficient (MCC), a widely used metric for imbalanced datasets. To our knowledge, very few studies have computed MCC scores to validate models for Mpox detection. Furthermore, no prior studies have conducted the comprehensive analysis that we present in this work.
iv. Finally, we make a comparison of our work with results published in prior studies.

We organize this paper as follows: In Section 2, we review existing studies on Mpox detection and highlight their research gaps. In Section 3, we discuss our dataset, train-test split, and data augmentation procedure and perform Kullback-Leibler divergence on both the original and augmented images. We also develop a hybrid deep learning framework in this section. Moreover, we provide an overview of the pre-trained DL and ML methods and metrics used in this study. In Section 4, we present the results and compare them with prior studies. Finally, we conclude the paper in Section 5.

## 2. Literature review

This section elaborates the introduction by reviewing existing techniques implemented in Mpox detection. Mpox is an infectious disease and one can study the dynamics of an infectious disease by developing models using a system of ordinary differential equations. A simple framework for these models is the SIR (susceptible, infectious, recovered) model, given by [28]:

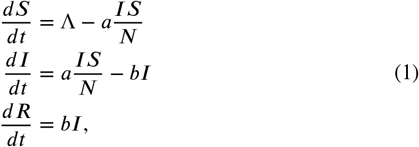

with initial conditions *S*(0) = *S*_0_, *I*(0) = *I*_0_, *R*(0) = *R*_0_. Here *S, I*, and *R* are the number of individuals in the susceptible, infectious, and recovered compartments, Λ represents the recruitment number in the susceptible compartment, *a* is the infection or disease transmission rate from susceptible to infection, and *b* denotes the recovery rate. Note that *t* > 0 and the total population is given by:

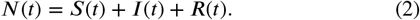

Using this model, one can compute the basic reproduction number ℛ _0_, which is a threshold that determines whether the disease will spread or die out. Due to the mathematical elegance, this model is widely popular and found wide ranging applications in the published literature. Recently, this infectious disease model has been studied to analyze the dynamics of Mpox and predict the virus’s spread [33, 3, 54].

[44] proposed a SEIR-type model for continuous monitoring and early intervention of the spread of Mpox virus. They proposed a robust model and performed sensitivity analyses for different parameters such as infection rate and recovery rate. Most importantly, they estimated the basic reproduction number for 2022 outbreak to investigate how rapidly the virus could spread.

[9] investigated the dynamics of Mpox outbreak using fractional-order SEIR model. They analyzed the effects of vaccination in the transmission and mitigation of Mpox virus. One can find a similar type of model outlined in [1].

[10] developed a model focusing on pair formation that reflects how close contact with individuals contributes to the spread of Mpox. Additionally, they investigated recovery dynamics to observe how quickly infected individuals recover and how this recovery rate affects the overall spread of the infection. Using infectious disease modeling and statistical techniques, [71] forecasted the global spread and trajectory of the Mpox virus. They emphasized identifying key factors responsible for the spread of MPXV. By analyzing the current data, they predicted the future scenario of the outbreak.

[36] performed a comprehensive analysis of the 2022 Mpox outbreak in New York City. They analyzed and compared different mathematical models, including SEIR, and evaluated their effectiveness in predicting and forecasting outbreak patterns. [48] has provided a comprehensive review on infectious disease modeling to investigate the Mpox transmission.

Although these models have received significant attention for investigating the dynamics of Mpox transmission, they are not well-suited for early disease prediction. Machine learning and deep learning techniques fill this gap.

[5] created an image dataset to detect and analyze Mpox with DL algorithms. The dataset consists of images with a resolution of 224×224×3 and allows the implementation of DL models. [5] applied three pretrained models, VGG-16, InceptionV3, and ResNet-50, to classify Mpox images. In addition to that, they combined these DL algorithms by employing majority voting and proposed an ensemble model. Among these models, ResNet-50 achieved 82.96% accuracy, 87% precision, 83% recall, and 84% F1 score, while other models, including the ensemble, produced lower scores in every metric. However, this study did not mention whether their data augmentation introduced any biases in their training dataset, which is clearly a significant research gap.

Using the same dataset, [58] developed a DL framework tailored for mobile-based applications to diagnose Mpox lesions. They obtained 91.11% accuracy, 90% precision, 90% recall, and 90% F1 score. This study did not validate the augmented images for their medical relevance.

[62] proposed an ensemble-based DL framework focused not only on Mpox detection but also on classifying other skin diseases. Their dataset was curated with samples of different skin diseases. Initially, they implemented 13 different DL techniques, then ensembled two best-performing algorithms—Xception and DenseNet-169 and achieved 87.13% accuracy, 85.44% precision, 85.47% recall, and 85.40% F1 score. However, this study analyzed their models only based on accuracy, precision, recall, and F1-scores. Note that imbalanced data could produce biased accuracy. Additionally, this study did not compute metrics such as the MCC score, which is widely used to evaluate model performance on imbalanced data.

[23] explored ResNet-50, EfficientNetb3, and EfficientNetb7 on a dataset that contains 160 Mpox lesions, 178 chickenpox, 54 cowpox, 358 small pox, and 50 healthy skins. They obtained 87% accuracy, 92% precision, 87% recall, and 90% F1 score by EfficientNetb3. Although this study trained several deep learning algorithms to detect Mpox, similar to prior studies ([5, 58]), they did not investigate whether their data augmentation introduced any biases. Additionally, they did not compute MCC scores or any other metrics tailored for imbalanced data.

[67] proposed a hybrid DL framework for Mpox detection. Unlike developing an ensemble model employing majority voting, they combined the two models with the highest accuracy with the Long Short-term Memory (LSTM) model and obtained 87% accuracy, 84% precision, 87% recall, and 85% F1 score. This study did not address the interpretability of the model’s decisions, which is considered a crucial component in medical diagnostics for clinical acceptance and trust.

[50] tested various deep learning models, including GoogleNet, ResNet-18, ResNet-50, ResNet-101, and SqueezeNet, for multi-class classification and Mpox detection, achieving an average accuracy of 91.19%. Explainable artificial intelligence (XAI) plays a crucial role in medical research, and [50] introduced XAI to make the deep learning model more interpretable. However, there is no discussion on the potential biases that could be introduced by the data augmentation process.

[47] used VGG19, Xception, InceptionV3 transfer learning models on augmented dataset created by [4], which includes, 1428 Mpox and 1764 others Skin lesions images. They achieved 98% accuracy by using InceptionV3 model. It is important to note that this study used augmented data created by [5]. As previously mentioned, there is no information on the qualitative or quantitative validation conducted to ensure that these augmented images do not introduce bias.

[70] explored vision transformers (VITs) and convolutional neural networks (CNNs) for Mpox lesion analysis. They proposed an ensemble learning strategy using the bagging ensemble technique and obtained 81.91% accuracy. This study did not explore different data augmentation strategies, nor did it address the interpretability of the Vision Transformer and CNN.

## 3. Methodology

In this section, we discuss our data, along with the train-test split and data augmentation procedure. Moreover, we introduce Kullback-Leibler divergence as a validation tool to demonstrate that augmentation does not significantly alter the data distribution. We also explain the hybrid DL framework for Mpox detection. Additionally, we briefly describe the DL architecture and ML classifiers, along with various evaluation metrics.

### 3.1. Data description

We use a publicly available dataset for Mpox detection, created by [5]. The dataset contains images with a resolution of 224 × 224 × 3. It is divided into two categories–Mpox and non-Mpox, where the latter includes images of other diseases–chickenpox and measles. In Table 1, we present a summary of the dataset, including the number of images in each category.

**Table 1.** Summary of the dataset.

| Category | Number of Images | Percentage |
| --- | --- | --- |
| Mpox | 102 | 45% |
| Others | 126 | 55% |
| **Total** | **228** | **100%** |

#### 3.1.1. Data preprocessing

In this step, we clean the data to ensure that there are no mislabeled or noisy images. Generally, image intensity values range from 0 to 255. We normalize the images so that the pixel values lie between 0 and 1.

#### 3.1.2. Train-test split

We split the dataset into two subsets—the train set, which contains 80%, and the test set, which includes 20% of the samples from the original dataset. We present a detailed summary in Table 2.

**Table 2.** Summary of the train and test data.

| Category | Training set | Test set |
| --- | --- | --- |
| Mpox | 81 | 21 |
| Others | 100 | 26 |
| **Total** | **181** | **47** |

### 3.2. Data augmentation

From Tables 1 and 2, we can see that the dataset is imbalanced, with significantly fewer Mpox images compared to other skin lesion images. To create robust models, we perform data augmentation on the training data. We employ various strategies, including rotation, horizontal flipping, width and height shifts, zooming, fill mode adjustments, and shear range modifications. Table 3 presents the total number of augmented images, while Table 4 provides detailed information on the augmentation strategies and their optimized values.

**Table 3.** Summary of the augmented images.

| Category | Augmented images |
| --- | --- |
| Mpox | 200 |
| Others | 200 |
| **Total** | **400** |

**Table 4.** Summary of the augmentation parameters.

| Augmentation Parameter | Description | Value |
| --- | --- | --- |
| Rotation range | Degree range for random rotation | 20 degree |
| Width shift range | Range of random horizontal shift | 20% of the image width |
| Height shift range | Range of random vertical shift | 20% of the image height |
| Shear range | Transformation that slants the image shape | 0.2 radians |
| Zoom range | Range of random zooming | 20% of the image |
| Horizontal flip | Randomly flip images horizontally | True |
| Fill mode | Fill strategy for newly created pixels | Nearest |

Note that each extracted feature is a tensor of dimension $l \times p \times q$ , where $l$ and $p$ are the height and width of the feature map and $q$ denotes the number of feature channels. In order to train a ML classifier, we have reshaped each of these features into a $d$ -dimensional vector where $d = l \times p \times q$ . The reshaping procedure is performed as follows:

#### 3.2.1. Kullback-Leibler divergence

Kullback-Leibler (KL) divergence, also known as relative entropy, is an essential metric for determining statistical differences between two distributions. We compute KL divergence to assess whether there is a significant difference between the original and augmented images. For empirical distributions *p* and *Q* defined on the same sample space 𝒱, KL divergence is defined as [19]:

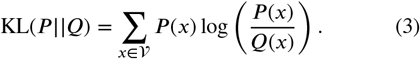

Equation (3) shows that the two distributions *p* and *Q* are nearly identical if KL(*P* ||*Q*) is zero or approximately close to zero.

### 3.3. Hybrid deep learning framework for Mpox detection

Consider the dataset 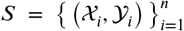, where 𝒳 _*i*_ denotes the *i*^*th*^ image and 𝒴_*i*_ represents its corresponding image label. We develop the framework by combining a pre-trained DL architecture and an ML classifier in the following steps:

Step 1 (Preprocessing): In this initial phase, we clean the dataset through various processes, including noise reduction, handling mislabeled images, and removing duplicate images to ensure the dataset is prepared for optimal model performance.

Step 2 (Train-test split): We partition the given dataset *S* into two distinct subsets, *S*_Train_ and *S*_Test_, such that

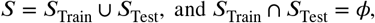

where *S*_Train_ denotes the training set containing 80% of the sample and *S*_Test_ denotes the test set containing 20% of the samples from the dataset *S*. We perform data augmentation on *S*_Train_.

Step 3 (Feature extraction): In this step, using a pre-trained DL architecture ℱ, we extract features from each image as follows:

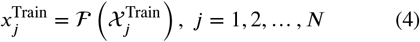

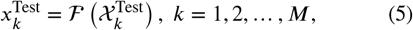

where 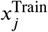 and 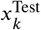 represent the features extracted from 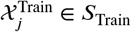 and 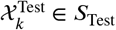 respectively.

Note that each extracted feature is a tensor of dimension *l*×*p*×*q*, where *l* and *p* are the height and width of the feature map and *q* denotes the number of feature channels. In order to train a ML classifier, we have reshaped each of these features into a *d*-dimensional vector where *d* = *l* × *p* × *q*. The reshaping procedure is performed as follows:

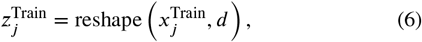

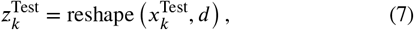

where *j* ranges from 1 to *N*, and *k* ranges from 1 to *M*. Step 4 (Train classification model): Using these features 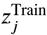, we train a classification model 𝒢. After that, we test the model using the features 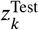 and predict the labels 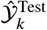 as follows:

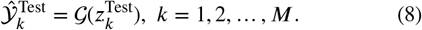

Step 5 (Model Evaluation): Finally, we evaluate the hybrid model using standard metrics such as accuracy, precision, weighted precision, recall, weighted recall, F1 score, weighted F1 score, and Matthews Correlation Coefficient (MCC) score. We discuss these metrics in subsection 3.6.

In Figure 2, we illustrate this framework in detail. Additionally, we include all essential variables and notations, along with their descriptions, in Table 5.

**Table 5.** Description of the notations and variables.

| Notations | Description |
| --- | --- |
| $S$ | Image dataset |
| $S_{\text{Train}}$ | Training dataset |
| $S_{\text{Test}}$ | Test dataset |
| $n$ | Total number of images |
| $N$ | Total number of images in $S_{\text{Train}}$ |
| $M$ | Total number of images in $S_{\text{Test}}$ |
| $\mathcal{F}$ | Pre-trained deep learning architecture |
| $\mathcal{G}$ | Classification model |
| $\mathcal{X}_i$ | $i^{\text{th}}$ image in $S$ |
| $\mathcal{Y}_i$ | Label of the $i^{\text{th}}$ image $\mathcal{X}_i$ |
| $\mathcal{X}_j^{\text{Train}}$ | $j^{\text{th}}$ image in $S_{\text{Train}}$ |
| $\mathcal{X}_k^{\text{Test}}$ | $k^{\text{th}}$ image in $S_{\text{Test}}$ |
| $x_j^{\text{Train}}$ | Extracted feature from $\mathcal{X}_j^{\text{Train}}$ |
| $x_k^{\text{Test}}$ | Extracted feature from $\mathcal{X}_k^{\text{Test}}$ |
| $l$ | Height of the feature map |
| $p$ | Width of the feature map |
| $q$ | Number of feature channels |
| $d$ | Dimension of the reshaped features |
| $z_j^{\text{Train}}$ | Reshaped training feature |
| $z_k^{\text{Test}}$ | Reshaped test feature |
| $\hat{\mathcal{Y}}$ | Predicted labels |
| $H_0$ | Null hypothesis |
| $H_A$ | Alternative hypothesis |
| TP | True positive |
| TN | True negative |
| FP | False positive |
| FN | False negative |

**Figure 2:**
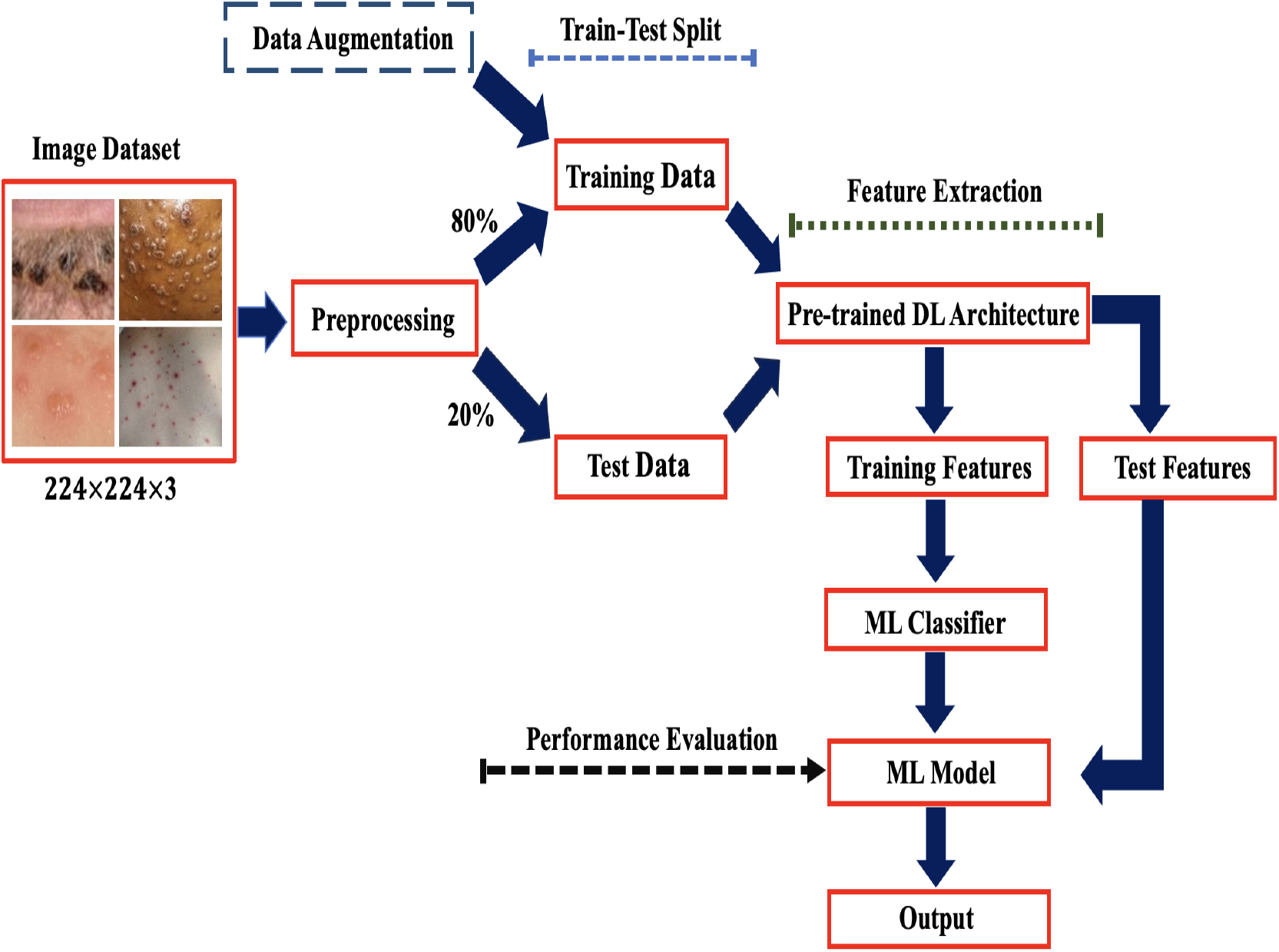
A hybrid deep learning framework for Mpox detection, combining a pre-trained deep learning architecture and machine learning classifier.

### 3.4. Pre-trained deep learning architectures

We implement several pre-trained DL architectures for feature extraction, including DenseNet, MobileNet, Inception, Inception-Residual Network, and Xception. We discuss them briefly in the following subsections.

#### 3.4.1. DenseNet

All the layers of this architecture are connected in a feedforward manner, where all the features from the preceding layers are concatenated and transferred to the successive layers [22]. This process allows to reuse the features, leverage the gradient flow, and prevent the vanishing gradient problem. In addition to that, it has transition layers that consist of batch normalization, ReLu activation function, 1 × 1 convolution to reduce dimensions, and 2 × 2 average pooling that controls the growth of the number of features and consequently reduces the computational complexity [22]. We implement three versions of DenseNet for feature extraction: DenseNet-121, DenseNet-169, and DenseNet-201, which contain 121, 169, and 201 layers, respectively.

#### 3.4.2. Inception

This DL architecture uses an inception module, which allows Inception to learn global and local patterns and leverage its ability to learn features comprehensively. More specifically, the inception module applies convolutional filters of different sizes (1 × 1, 3 × 3, and 5 × 5) in parallel, which allows this architecture to learn features simultaneously at multiple scales [6]. In addition, 1 × 1 convolution reduces the dimensionality before applying the larger filters, which leverages the architecture’s computational efficiency [6]. In this paper, we use InceptionV3 for feature extraction.

#### 3.4.3. Inception-residual network

It ensembles the architecture of Inception and residual networks, incorporating residual connections within the inception block structure. Residual connections resolve the vanishing gradient issue and enhance the efficiency of this architecture in classification tasks, particularly for image data [6]. We use the Inception-ResNetV2 architecture for feature extraction in this paper.

#### 3.4.4. Xception

This architecture is an improvement over Inception. One can consider this a bundle of depthwise separable convolution (DSC) with residual connection (RC), where DSC reduces the computational cost as well as improves memory usage by separating the learning of channel-wise and space-wise features, and RC solves the vanishing gradient issue [6].

#### 3.4.5. MobileNet

It is an alternative to CNNs, computationally more efficient than CNNs and tailored for deployment on devices with limited computational capacities, such as smartphones [29]. Unlike CNNs, which use regular convolution operations where a filter processes all the input channels (e.g., red, green, blue for a color image) together, MobileNet uses a depthwise separable convolution operation [51]. This convolution operation comprises into two steps— Depthwise convolution (DC) and Pointwise convolution (PC) [51]. Initially DC, filters are applied to each input channel individually and reduce the number of parameters, which makes the computational process faster compared to the regular convolution operation. Then, PC combines the outputs from DC and transforms them into a single output [51]. In this paper, we implement MobileNetV1 and MobileNetV2 for feature extraction from images.

We utilize Python packages from the scikit-learn library to implement these deep learning architectures [55]. These models utilize the ReLU (Rectified Linear Unit) activation function, defined as:

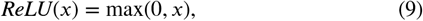

and are trained on the ImageNet database. We provide a summary of these deep learning architectures in Table 6.

**Table 6.** Summary of the deep learning architectures.

| Architectures | Activation function | Weights | Input image shape | Key Differences |
| --- | --- | --- | --- | --- |
| DenseNet-201 | ReLU | ImageNet | $224 \times 224 \times 3$ | Consists of 201 layers |
| DenseNet-169 | ReLU | ImageNet | $224 \times 224 \times 3$ | 4 dense blocks with 169 layers |
| DenseNet-121 | ReLU | ImageNet | $224 \times 224 \times 3$ | 4 dense blocks with 121 layers |
| InceptionV3 | ReLU | ImageNet | $224 \times 224 \times 3$ | Uses kernels of different sizes |
| InceptionResNetV2 | ReLU | ImageNet | $224 \times 224 \times 3$ | Includes residual connections |
| Xception | ReLU | ImageNet | $224 \times 224 \times 3$ | Applies depthwise separable convolutions |
| MobileNetV1 | ReLU | ImageNet | $224 \times 224 \times 3$ | Uses depthwise and pointwise convolutions |
| MobileNetV2 | ReLU | ImageNet | $224 \times 224 \times 3$ | Linear bottlenecks exist between layers |

### 3.5. Machine learning classifiers

We have used the following feature set

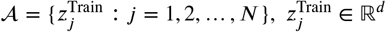

to train several ML classifiers. In the following subsections, we provide an overview of how these classifiers generally function.

#### 3.5.1. Support vector machine

A crucial step in training a support vector machine (SVM) is to find a hyperplane, given by:

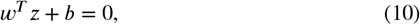

so that the margin between the nearest data points of each class and the hyperplane is maximized. Here, *w* ∈ ℝ^*d*^ denotes the weight, and *b* ∈ ℝ denotes the bias in (10). It is important to note that to train an SVM; the labels must be mapped either −1 and 1 or 0 and 1 [63, 65]. For linearly separable data, to obtain the final classifier *x* ↦ sign (*w* ^*T*^ *z* + *b*), one has to solve the following optimization problem [38]:

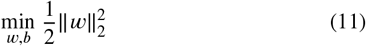

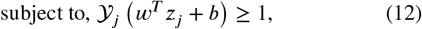

where *j* ∈ {1, 2, …, *N*}. Note that sign (*w*^*T*^ *z* + *b*) = 1 if *w*^*T*^ *z* + *b* > 0, and 0 otherwise.

In this paper, to train SVM, we have mapped the labels associated with each feature into 0 and 1.

#### 3.5.2. Random forest

It performs classification task by ensembling predictions made by multiple decision trees based on majority voting strategy [12]. Suppose there are 𝓁 decision trees *T*_1_, *T*_2_, …, *T*_𝓁_ in the forest. These trees are also known as “base learner” [20]. Initially, it trains each decision tree on a subset drawn from the training feature space using bootstrap sampling with replacement. Each of these trees make a prediction for a given input *z*, then final prediction is made as [20]:

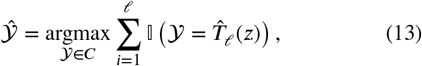

where 𝕀 denotes the indicator function, 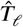 denotes the 𝓁-th fitted tree at *z*, and *C* is the set of all possible values of 𝒴.

#### 3.5.3. Logistic regression

For any probability of success *p* expressed in terms of odds as [41, 52]:

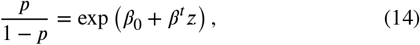

the Logisitic regression (LR) is defined as follows:

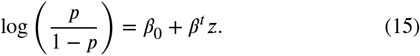

Here *β* = {*β*_1_, *β*_2_, …, *β*_*n*_} is the coefficient vector of *z* and *β*_0_ is a constant.

#### 3.5.4. Decision tree

In general, a Decision tree (DT) is trained by deploying a continuously growing binary decision tree in training features 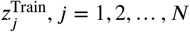 It starts from a root node and ends with the decision outputs known as the leaf node [21, 64].

#### 3.5.5. K-nearest neighborhood

The K-Nearest Neighbor (K-NN) classifier predicts the label of a target point *z*^*∗*^ based on the patterns that are nearest to *z*^*∗*^. To identify these patterns, the classifier computes the similarity between the target point *z*^*∗*^ and the input features using distance metrics such as the Euclidean distance or the Minkowski distance [57, 42]. For binary classification, K-NN is defined as [42]:

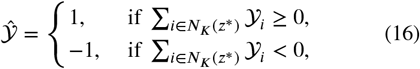

where *K* denotes the neighborhood size, and *N*_*K*_ (*z*^*∗*^) denotes the set of indices of the *K* nearest neighbor. In this paper, we consider *K* = 5.

#### 3.5.6. Naïve bayes classifier

It assumes that the features are conditionally independent given any target label [66]. In other words, it considers the contribution of each feature independently. For binary classification, it predicts a label as follows [66]:

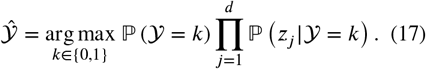

In this paper, for each *z*_j_, we have considered that the likelihood function ℙ (*z*_*j*_ |𝒴 =*k*) is normally distributed with mean μ_*jk*_ and variance *σ*_*i*_.

#### 3.5.7. Adaptive boosting

Adaptive boosting (AdaBoost) combines weak classifiers *T*_1_, *T*_2_, …, *T*_*m*_ and creates a strong classifier as follows:

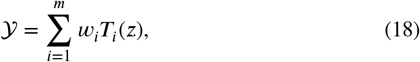

where *m* denotes the total number of classifiers, and *w*_*i*_ is the weight assigned to the weak classifier *T*_*i*_ (see[26]).

#### 3.5.8. Extreme gradient boosting (XGBoost)

It is an improvement over Gradient boosting (GB), which forms a model for prediction by ensembling multiple decision trees where each successive tree depends upon the previous tree and tries to fix errors made by the preceding ones [17]. One of the important features of XGBoost is it prevents overfitting by regularizing the loss function as follows:

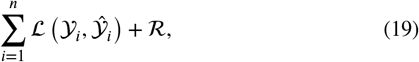

where 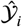 denotes the predicted observations, ℒ denotes the loss ^*i*^function, and ℛ is the regularization term, given by:

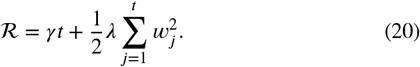

Here λ denotes the regularized parameter to scale the penalty, *γ* the minimum loss needed to further partition the leaf node, *w* denotes the weight of the leafs, *t* is the number of leaves in the tree.

#### 3.5.9. Light gradient boosting (LightGBM)

It was built at the top of the Gradient boosting framework for classification and other machine-learning tasks. It was developed by Microsoft [37]. Its architecture includes histogram based learning, which reduces memory usage and accelerates computational speed, leaf-wise tree growth, Gradient based one-sided sampling (GOSS), exclusive feature bundling, which groups categorical features with common values, parallel and GPU learning, and regularization to prevent overfitting such as *L*_1_ or *L*_2_ regularization [37].

Besides AdaBoost, XGBoost, and LightGBM, there are other gradient boosting methods, such as CatBoost, which we found to be more time-consuming to train compared to the others. As a result, we explored computationally efficient gradient boosting classifiers. We implemented these classifiers using Python packages and modules provided by the Scikit-learn library [55]. We summarize the hyperparameters of these classifiers, along with their optimal values, in Table 7. We optimized these hyperparameters through the grid search algorithm.

**Table 7.** Summary of hyperparameters and optimal values for machine learning classification models.

| Models | Tuning Parameters | Best Parameters | Description |
| --- | --- | --- | --- |
| SVM | $C = [10^{-1}, 10^3]$,<br>$\gamma = [10^{-4}, 1]$,<br>Kernel = 'Linear' | $C = 0.1$,<br>$\gamma = 1.0$ | $C$, regularization parameter.<br>$\gamma$ controls a training example's influence. |
| RF | $\mu = \{3, 5, 10, \infty\}$,<br>$n = \{10, 100, 200\}$,<br>$\alpha = \{1, 3, 5, 7\}$,<br>$\ell = \{1, 2, 3\}$,<br>$m = \{2, 3\}$ | $\mu = 3$,<br>$n = 100$,<br>$\alpha = 5$,<br>$\ell = 1$,<br>$m = 3$ | $\mu$, maximum depth of the decision trees.<br>$n$, number of trees in the forest.<br>$\alpha$ limits the features used for node splits.<br>$\ell$ sets the minimum samples per leaf.<br>$m$ sets the minimum samples to split a node. |
| LR | $C = [10^{-3}, 10^3]$,<br>Penalty $\in \{\ell_1, \ell_2\}$ | $C = 0.1$,<br>Penalty = $\ell_2$ | $C$, regularization parameter.<br>Penalty, specifies the type of regularization applied. |
| DT | $f = \{'auto', 'sqrt', 'log2'\}$,<br>$\bar{\alpha} = [10^{-2}, 10^{-1}]$,<br>$\mu = \{5, 6, 7, 8, 9\}$,<br>$C = \{'gini', 'entropy'\}$ | $f = 'log2'$,<br>$\bar{\alpha} = 0.01$,<br>$\mu = 7$,<br>$C = 'entropy'$ | $f$, number of features to consider when splitting a node.<br>$\bar{\alpha}$, controls tree pruning.<br>$\mu$, maximum depth of the tree.<br>$C$, function to measure the quality of a split. |
| K-NN | $N = \{5, 7, 9, 11, 13, 15\}$,<br>$W = \{'uniform', 'distance'\}$ | $N = 5$,<br>$W = 'uniform'$ | $N$, number of nearest neighbors.<br>$W$ denotes weight. |
| NB | $\delta = \text{LogSpace}[10^{-9}, 1]$ | $\delta = 0.01874$ | $\delta$, smoothing parameter. |
| XGBoost | $\mu = \{3, 5, 7\}$,<br>$r = \{0.1, 0.01, 0.001\}$,<br>$J = \{0.5, 0.7, 1\}$ | $\mu = 3$,<br>$r = 0.1$,<br>$J = 0.5$ | $\mu$, maximum depth.<br>$r$, learning rate.<br>$J$, fraction of the training data to use for each boosting round. |
| AdaBoost | $n = \{50, 100, 150\}$,<br>$r = \{0.01, 0.1, 1.0\}$ | $n = 3$,<br>$r = 0.1$ | $n$, number of trees in the forest.<br>$r$, learning rate. |
| LightGBM | $\lambda = \{31, 63, 127\}$,<br>$\mu = \{-1, 3, 5\}$,<br>$J = \{0.8, 1.0\}$,<br>$\eta = \{0.8, 1.0\}$ | $\lambda = 31$,<br>$\mu = 3$,<br>$J = 0.8$,<br>$\eta = 0.8$ | $\lambda$, number of leaves.<br>$\mu$, maximum depth.<br>$J$, fraction of the training data to use for each boosting round.<br>$\eta$, fraction of features to be randomly selected for each tree |

### 3.6. Evaluation metrics

In this section, we present evaluation metrics to evaluate hybrid DL models. Our goal is to classify Mpox and other conditions from image data. To formalize the evaluation process, we construct the following hypotheses:

*H*_0_ : The observation belongs to the “Others” class

*H*_*A*_ : The observation belongs to the “Mpox” class where *H*_0_ denotes the null hypothesis and *H*_*A*_ is the alternative hypothesis. Based on the predictions made by a hybrid DL model, the possible outcomes of this hypothesis test are:

i. True positive (TP): observation is correctly predicted as Mpox. In other words, *H*_0_ is rejected.
ii. True negative (TN): *H*_0_ is accepted, that is observation is classified as others.
iii. False positive (FP): observation is incorrectly classified as Mpox when it is actually others, which means that the *H*_0_ is rejected when it is true.
iv. False negative (FN): observation is incorrectly classified as others when it is actually Mpox. In other words, *H*_0_ is accepted when it is false.

These outcomes can be represented in a matrix form, commonly referred to as the confusion matrix (CM), as follows:

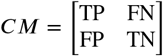

We have computed accuracy, precision, recall, F1 score, weighted precision, weighted recall, weighted F1 score, and Matthews Correlation Coefficient (MCC) using these outcomes. In Table 8, we present the evaluation metrics in detail.

**Table 8.** Evaluation metrics.

| Metric | Formula |
| --- | --- |
| Accuracy | $\frac{TP+TN}{TP+TN+FP+FN}$ |
| Precision | $\frac{TP}{TP+FP}$ |
| Recall (Sensitivity) | $\frac{TP}{TP+FN}$ |
| F1 Score | $2 \times \frac{\text{Precision} \times \text{Recall}}{\text{Precision} + \text{Recall}}$ |
| Weighted Precision | $\frac{\sum_{i=1}^k w_i \times \text{Precision}_i}{\sum_{i=1}^k w_i}$ |
| Weighted Recall | $\frac{\sum_{i=1}^k w_i \times \text{Recall}_i}{\sum_{i=1}^k w_i}$ |
| Weighted F1 Score | $\frac{\sum_{i=1}^k w_i \times F1_i}{\sum_{i=1}^k w_i}$ |
| MCC score | $\frac{(TP \times TN) - (FP \times FN)}{\sqrt{(TP+FP)(TP+FN)(TN+FP)(TN+FN)}}$ |

In addition to these evaluation metrics, we have also computed the area under the receiver operating characteristic curve (ROC). ROC plots the sensitivity (true positive rate) against the false positive rate (FPR). FPR is given by:

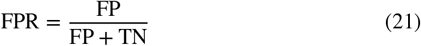

For *n* points on the ROC curve, area under the curve (AUC) can be computed using Trapezoidal rule approximation as follows:

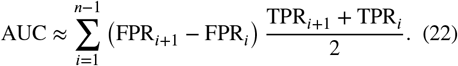

## 4. Results and discussion

Lack of skin lesion images and computational complexity are two major challenges in developing deep learning frameworks for Mpox detection. The objective of our hybrid deep learning framework is to detect Mpox using a small dataset while optimizing memory usage and computational time.

As an initial step to ensure model robustness, we performed data augmentation in the training dataset *S*_Train_. To address that data augmentation is not going to introduce any bias, we performed KL divergence test between original and augmented Mpox images as well as original and augmented other images. We present these results in Figure 3. Furthermore, we have comprehensively analyzed this framework using several hybrid DL models (see Table 9, Table 10, and Table 11). The hybrid models listed in Table 9 combine differ-DenseNet architectures (DenseNet-201, DenseNet-169, and DensNet-121) with ML classifiers. Notably, the hybrid model consists of DenseNet-201, and LR (D201LR) obtained an accuracy of 85.11%, outperforming other models that incorporated DenseNet-201 as a feature extraction tool. This model identified 18 true positive Mpox images out of 21 Mpox cases reported in the test dataset (see Figure 4a). On the other hand, DenseNet-201 combined with K-NN (D201KNN) achieved the highest precision (90%) among all DenseNet-201 incorporated hybrid models, highlighting its ability to minimize false positive cases.

**Table 9.**
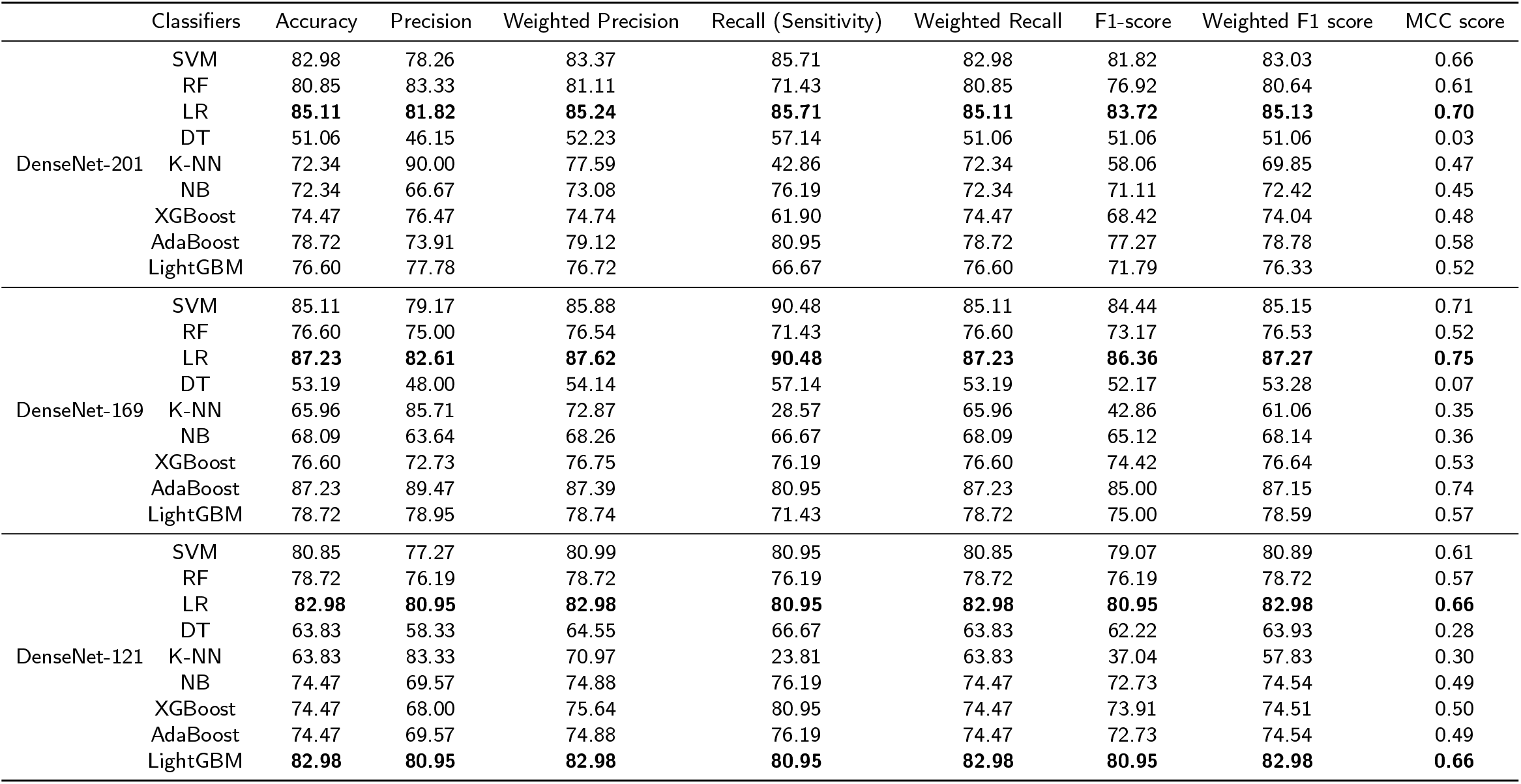
Hybrid DL models combining DenseNet architectures and ML Classifiers.

**Table 10.** Hybrid DL models combining InceptionV3, InceptionResNetV2, and Xception architectures with ML classifiers.

|  | Classifiers | Accuracy | Precision | Weighted Precision | Recall (Sensitivity) | Weighted Recall | F1-score | Weighted F1 score | MCC score |
| --- | --- | --- | --- | --- | --- | --- | --- | --- | --- |
| InceptionV3 | SVM | 80.85 | 77.27 | 80.99 | 80.95 | 80.85 | 79.07 | 80.89 | 0.61 |
|  | RF | 82.98 | 78.26 | 83.37 | 85.71 | 82.98 | 81.82 | 83.03 | 0.66 |
|  | LR | 82.98 | 80.95 | 82.98 | 80.95 | 82.98 | 80.95 | 82.98 | 0.66 |
|  | DT | 78.72 | 73.91 | 79.12 | 80.95 | 78.72 | 77.27 | 78.78 | 0.58 |
|  | K-NN | 63.83 | 70.00 | 65.66 | 33.33 | 63.83 | 45.16 | 60.57 | 0.26 |
|  | NB | 74.47 | 71.43 | 74.47 | 71.43 | 74.47 | 71.43 | 74.47 | 0.48 |
|  | XGBoost | 85.11 | 76.92 | 87.05 | 95.24 | 85.11 | 85.11 | 85.11 | 0.72 |
|  | AdaBoost | **87.23** | **82.61** | **87.62** | **90.48** | **87.23** | **86.36** | **87.27** | **0.75** |
|  | LightGBM | 82.98 | 80.95 | 82.98 | 80.95 | 82.98 | 80.95 | 82.98 | 0.66 |
| InceptionResNetV2 | SVM | 78.72 | 82.35 | 79.21 | 66.67 | 78.72 | 73.68 | 78.36 | 0.57 |
|  | RF | **85.11** | **88.89** | **85.50** | **76.19** | **85.11** | **82.05** | **84.94** | **0.70** |
|  | LR | 78.72 | 82.35 | 79.21 | 66.67 | 78.72 | 73.68 | 78.36 | 0.57 |
|  | DT | 68.08 | 66.67 | 67.94 | 57.14 | 68.09 | 61.54 | 67.73 | 0.35 |
|  | K-NN | 55.32 | 50.00 | 53.37 | 14.29 | 55.32 | 22.22 | 47.91 | 0.04 |
|  | NB | 68.09 | 66.67 | 67.94 | 57.14 | 68.09 | 61.54 | 67.73 | 0.35 |
|  | XGBoost | 78.72 | 73.91 | 79.12 | 80.95 | 78.72 | 77.27 | 78.78 | 0.58 |
|  | AdaBoost | 76.60 | 77.78 | 76.72 | 66.67 | 76.60 | 71.79 | 76.33 | 0.52 |
|  | LightGBM | 76.60 | 77.78 | 76.72 | 66.67 | 76.60 | 71.79 | 76.33 | 0.52 |
| Xception | SVM | **78.72** | **86.67** | **80.21** | **61.90** | **78.72** | **72.22** | **78.05** | **0.58** |
|  | RF | 70.21 | 73.33 | 70.80 | 52.38 | 70.21 | 61.11 | 69.27 | 0.39 |
|  | LR | **78.72** | **86.67** | **80.21** | **61.90** | **78.72** | **72.22** | **78.05** | **0.58** |
|  | DT | 65.96 | 63.16 | 65.76 | 57.14 | 65.96 | 60.00 | 65.74 | 0.31 |
|  | K-NN | 59.57 | 60.00 | 59.70 | 28.57 | 59.57 | 38.71 | 55.93 | 0.16 |
|  | NB | 65.96 | 57.14 | 76.24 | 95.24 | 65.96 | 71.43 | 63.94 | 0.43 |
|  | XGBoost | 70.21 | 73.33 | 70.80 | 52.38 | 70.21 | 61.11 | 69.27 | 0.39 |
|  | AdaBoost | 76.60 | 81.25 | 77.35 | 61.90 | 76.60 | 70.27 | 76.04 | 0.53 |
|  | LightGBM | 76.60 | 81.25 | 77.35 | 61.90 | 76.60 | 70.27 | 76.04 | 0.53 |

**Table 11.**
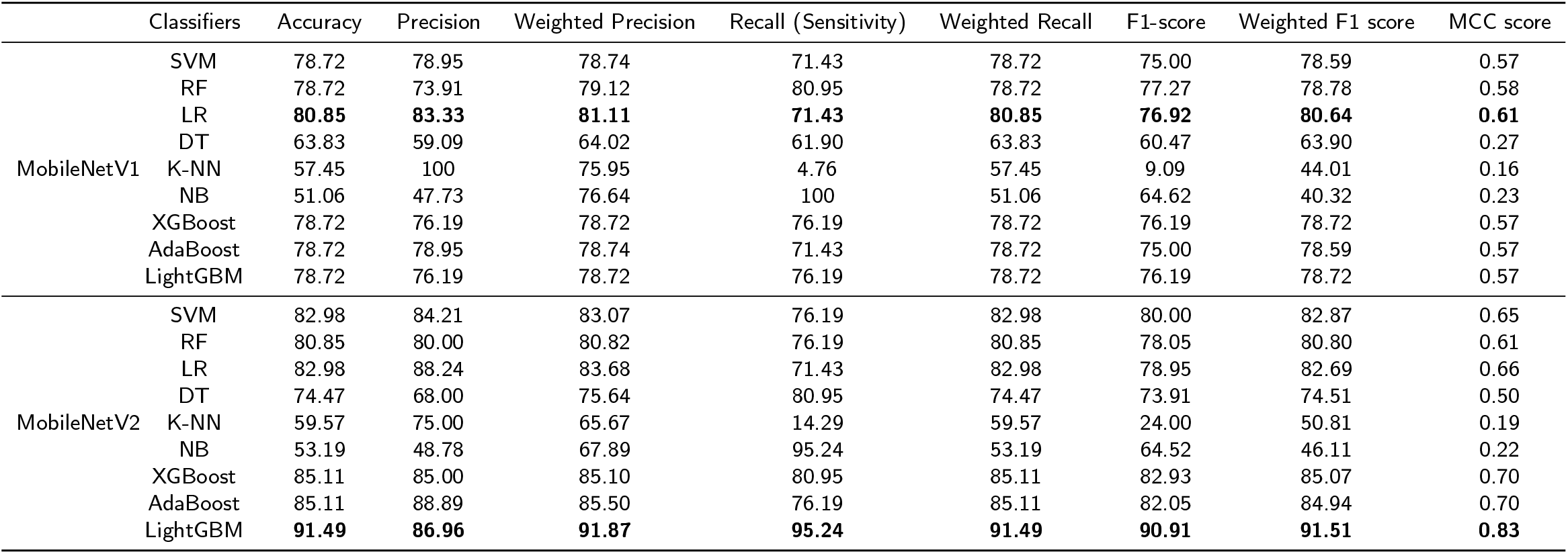
Hybrid DL models combining MobileNetV1 and MobileNetV2 architectures with ML classifiers.

**Figure 3:**
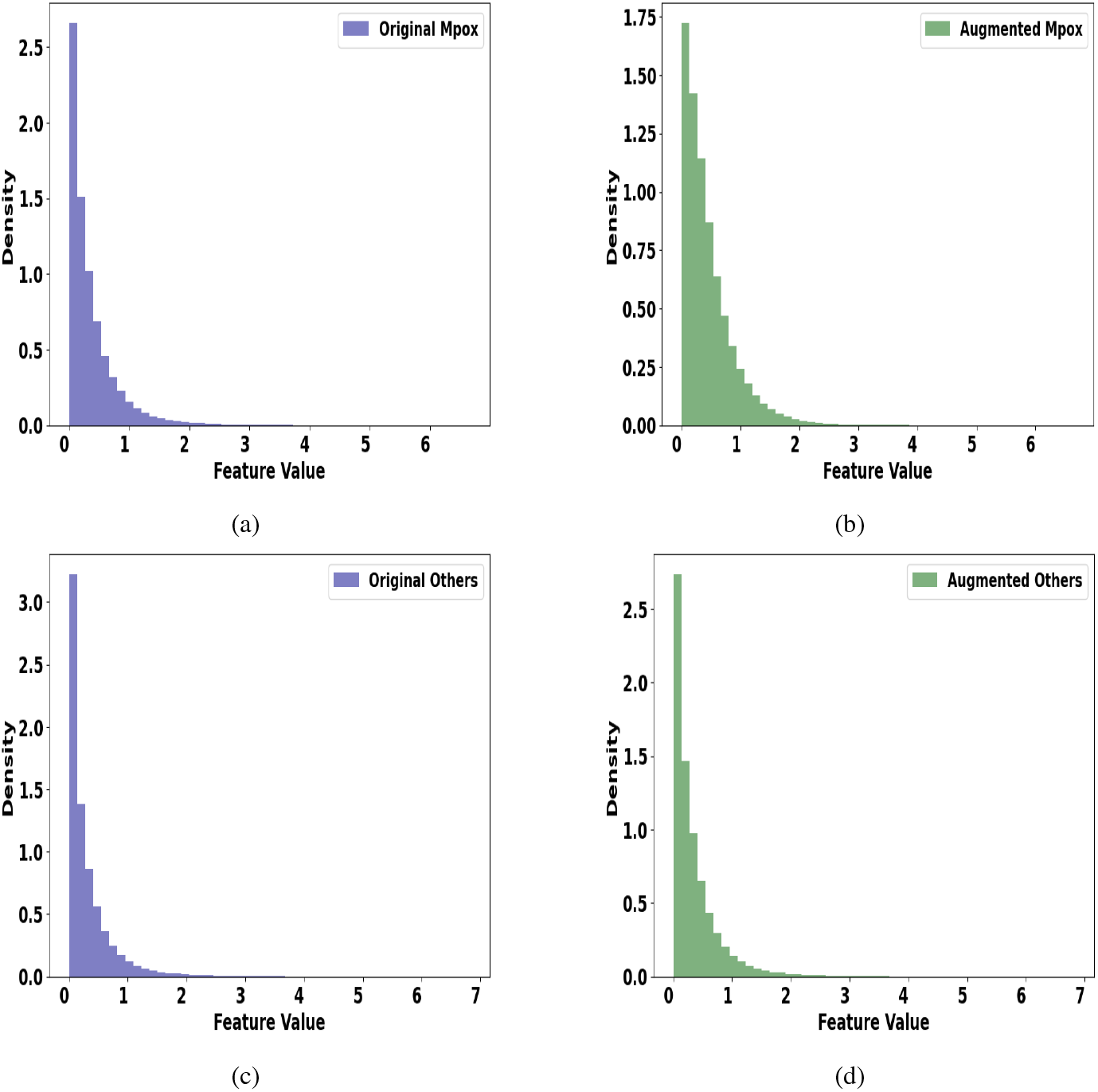
The Kullback-Leibler (KL) divergence analysis compares the distributions of original and augmented images. Figures 3a and 3b depict the distribution of original and augmented Mpox images. Both of these two figures exhibit a right-skewed distribution, indicating that augmentation has preserved the original data distribution. The KL divergence score for Mpox images is 0.0505, indicating no significant difference between the distributions. Similarly, Figures 3c and 3d illustrate the distribution of other images, with a KL divergence score of 0.0101, confirming that the augmentation process did not introduce any statistically significant differences.

**Figure 4:**
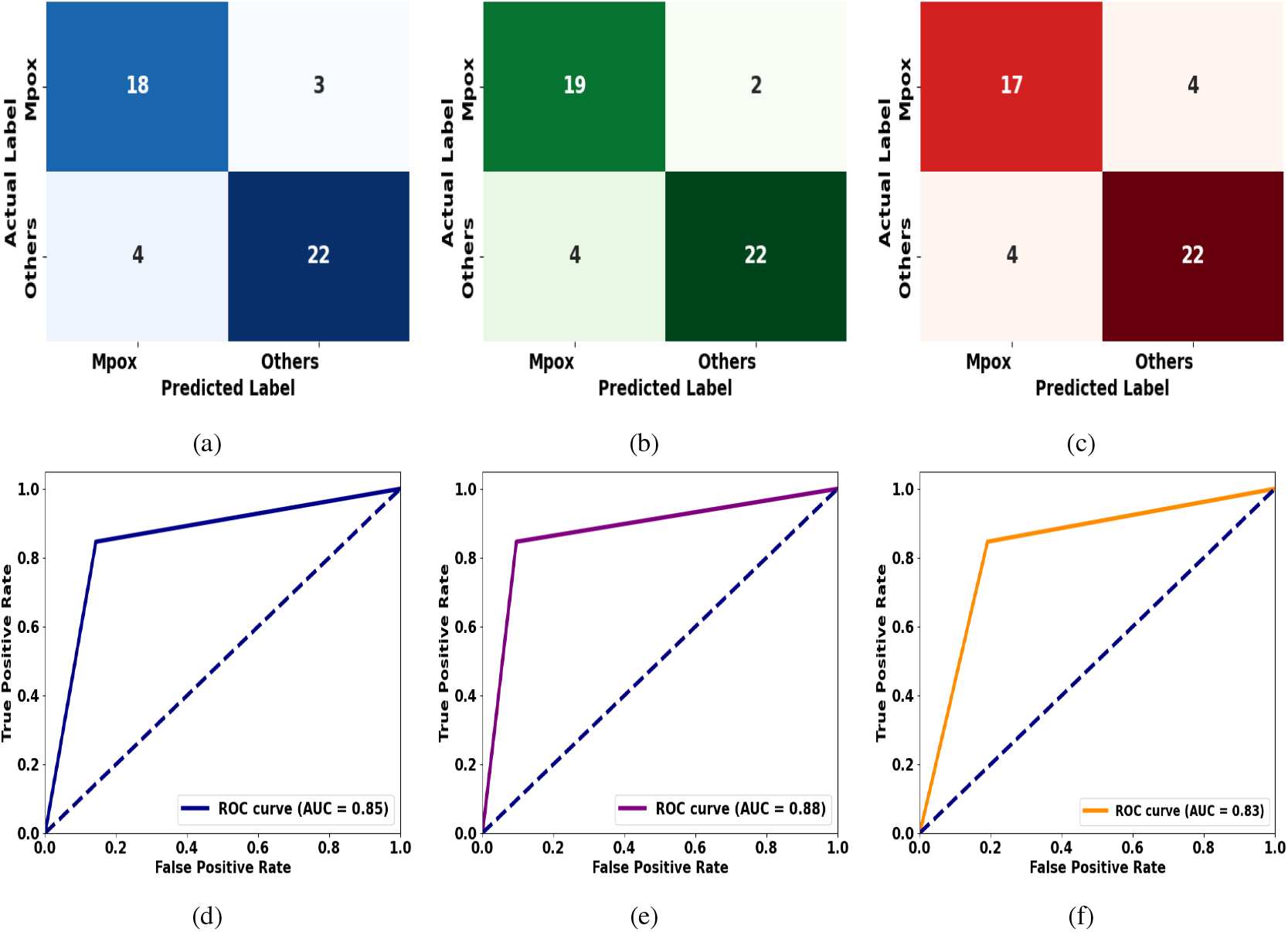
Confusion matrices and ROC curves for the selected models, incorporating DenseNet-201, DenseNet-169, and DenseNet-121 deep learning architectures as feature extraction tools from the image data. These models are selected based on their MCC score from each category listed in Table 9. Figure 4a, 4b and 4c are the confusion matrices for D201LR, D169LR, and D121LightGBM respectively, and 4d, 4e, and 4f illustrate their corresponding ROC curves. Confusion matrices display the counts of true positives, false negatives, false positives, and true negatives identified by these models. AUC scores measure the overall performance of these models in distinguishing between positive and negative classes by calculating the area under the ROC curve. A higher AUC indicates better discrimination between the two classes.

The highest overall accuracy, 87 23%, was achieved by the model consisting of DenseNet-169 with LR (D169LR). While it outperforms other hybrid models, its precision is significantly small (82.61%) compared to D201KNN. It shows that D169LR had a slightly higher rate of predicting false positive cases.

DenseNet-169 combined with ML classifier Adaboost (D169Adaboost) is another model which attained the same accuracy as D169LR (87.23%). Other evaluation metrics of this model, such as weighted precision (87.39%), weighted recall (87.23%), F1-score (85%), and weighted F1-score (87.15%) are comparable to D169LR, although its precision 89.47% is higher than D169LR (82.61 %), which indicates its strength while minimizing false positive cases is essential.

DenseNet-169 integrating with SVM (D169SVM) another noteworthy hybrid model for its highest recall rate, 90.48%, the same as D169LR. Hybird Models,

DenseNet-121 with LR (D121LR) and DenseNet-121 with LightGBM (D121LightGBM) both performed equally in terms of every metric for Mpox prediction (see Table 9).

However, D169LR outperformed other models in MCC score (Table 9). Note that, MCC score provides valuable information about the model performance, most importantly when the dataset is imbalanced. D169LR provided a score of 0.75, which highlights its strength in Mpox prediction. While D169Adaboost achieved a very close sore (0.74) to D169LR, performance of other competing models are significantly low. From the models enlisted in Table 9, we have selected three best performing models based on their MCC scores.

In Figure 4, we incorporated confusion matrices and ROC curves of these models. Since D121SVM and D121LightGBM performed equally in each evaluation metric, we only considered one of these two models in Figure 4. Confusion matrices allowed us to distinguish between positive and negative cases with accuracy. For example, from figure 4b, one can see that D169LR has predicted 19 true Mpox images which are clearly better than D201LR and D121lightGBM. Along with these metrics, we investigated ROC curves for these models to assess model performance.

AUC of D161LR reflects its efficiency in Mpox identification. AUC of D161LR is 0.88, which indicates that the model has an 88% probability of ranking a randomly chosen positive instance higher than a randomly chosen negative instance. In comparison, D201LR has an AUC of 0.85, and D121LightGBM has an AUC of 0.83, both of which are significantly lower than D161LR.

Although DenseNet architectures are very efficient feature extraction tools, these are computationally demanding in terms of computational resources. Therefore, we further investigated other DL architectures such as InceptionV3, InceptionResNetV2, Xception, MobileNetV1, and MobileNetV2.

In Table 10, we have constructed a few more hybrid models by integrating InceptionV3, InceptionRes-NetV2, and Xception architecture as a feature extraction tool. Out of all the models developed based on InceptionV3, InceptionV3 with AdaBoost (IV3AdaBoost) obtained the highest accuracy (87.23%). Although IV3XGBoost (InceptionV3 with XGBoost) outperformed IV3AdaBoost in sensitivity (95.24%), its MCC score (0.72) and other metric scores are significantly lower than IV3AdaBoost. IV3AdaBoost and D169LR performed equally to classify Mpox and other images since both models scored equal values in every evaluation metric.

InceptionResNetV2 with RF (IRV2LR) performed better in every metric compared to other models, which are based on InceptionResNetV2 architecture. Even though IRV2LR and D201LR have equal accuracy (85.11%) and MCC score (0.70), their precision, weighted precision, recall, weighted recall, F1 score, and weighted F1 score are significantly different.

However, AUC of D201LR (0.85) is slightly higher than IRV2LR (0.84). Among hybrid models integrating Xception as a feature extraction tool, XcepSVM (Xception with SVM) and XecpLR (Xeception with LR) performed better than others. However, its accuracy (78.72%) is significantly low compared to D169LR and IRV2LR. XcepSVM and XecpLR identified 13 images as Mpox (see Figure 5c).

**Figure 5:**
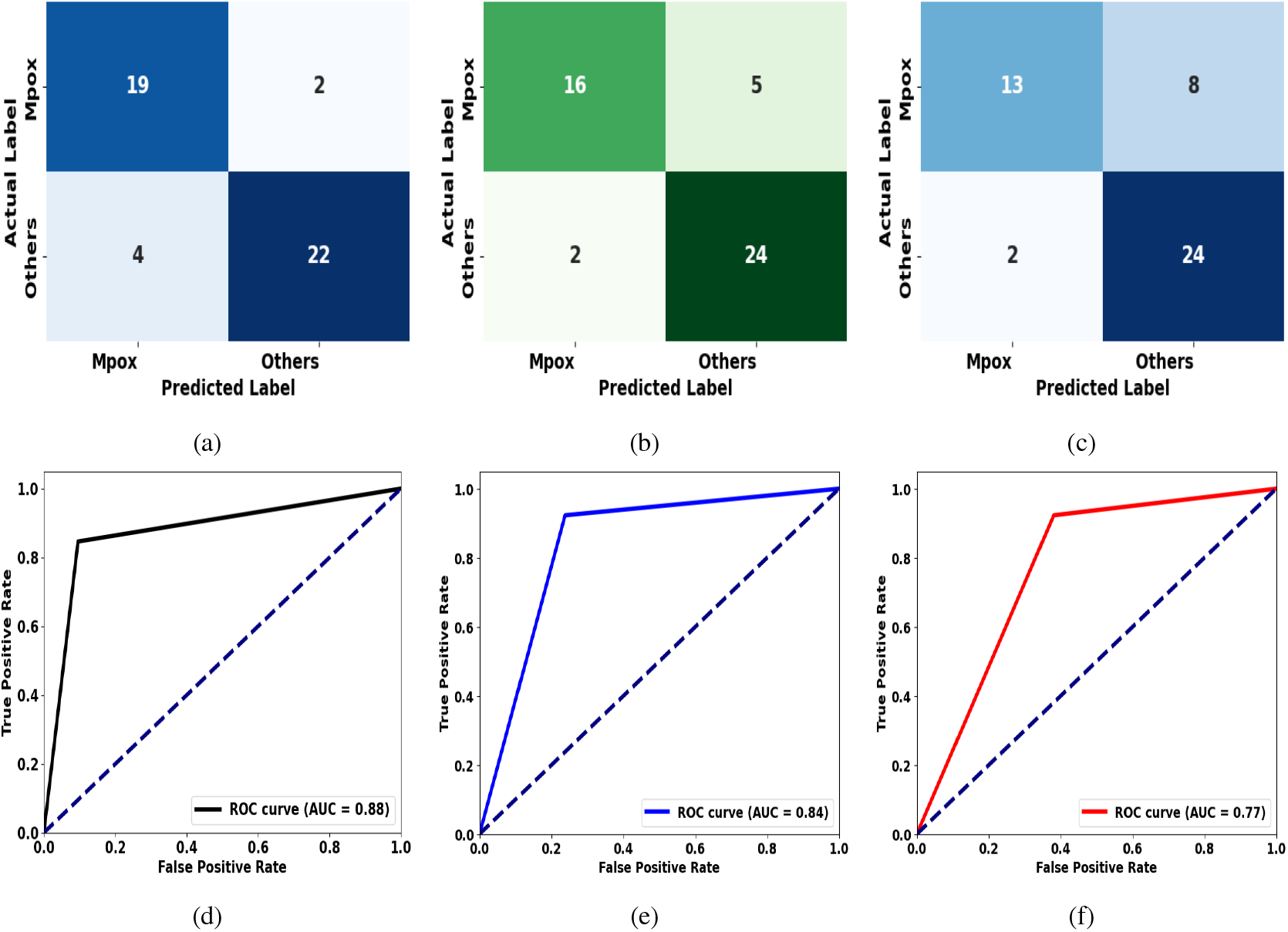
Performance evaluation of the models IV3AdaBoost (5a, 5d), IRV2LR (5b, 5e), and XcepSVM (5c, 5f) based on confusion matrices and ROC curves. These models are selected based on their MCC score from the list of models generated incorporating InceptionV3, InceptionResNetV2, and Xception as feature extraction tools (see Table 10). The AUC score and the number of true positive images identified by IV3AdaBoost highlight its efficiency in Mpox detection compared to the other two models.

Moreover, AUC of IV3AdaBoost is 0.88, which indicates that this model has 88% probability of identifying randomly chosen positive cases, which is 4% higher than IRV2LR and 11% higher than XcepSVM (see Figures 5d, 5e, and 5f). Since XcepSVM and XcepLR performed equally, we incorporated only the confusion matrix and ROC curve for XcepSVM (Figure 5c and5f). In Table 11, we have more hybrid models that were developed by combining MobileNet architectures and ML classifiers. MobileNetV1 and MobileNetV2 were used to create these models. MV1LR (MobileNetV1 with LR) obtained 80.85% accuracy, which indicates its efficiency in Mpox prediction. In addition to that, its other evaluation metrics, such as precision (83.33%), are also significantly higher than other models that were developed based on MobileNetV1. Although its sensitivity or recall (71.43%) is lower than the model MV1LightGBM (MobileNetV1 with LightGBM), which is 76.19%, its MCC score of 0.61 outperformed other models. It identified 15 Mpox images and 23 other images (see Figure 6a).

**Figure 6:**
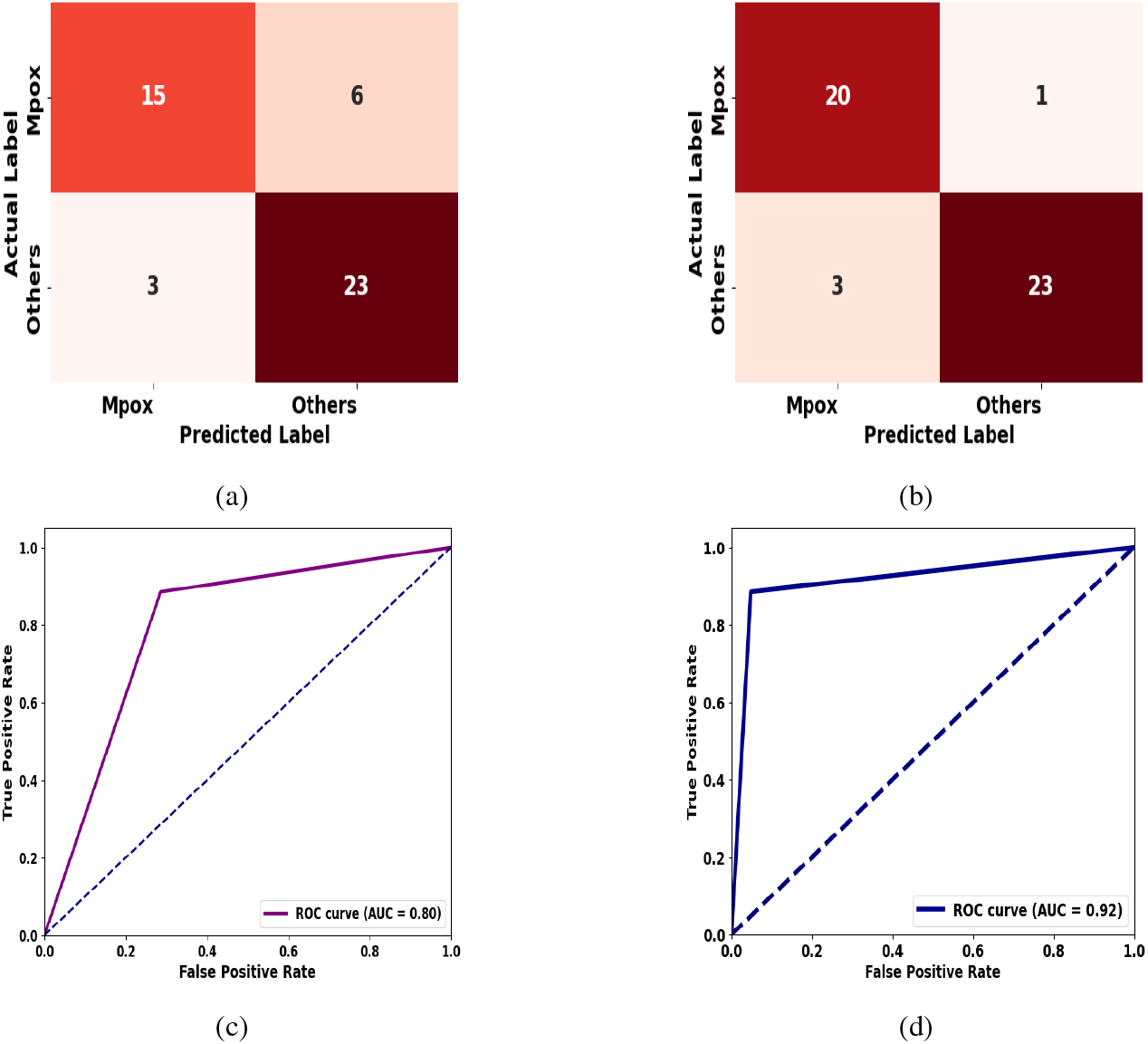
Comparison of the models constructed using MobileNetV1 and MobileNetV2 architectures. Figures 6a and 6c are the confusion matrix and ROC curve for MV1LR, respectively, while Figures 6b and 6d show the confusion matrix and ROC curve for MV2LightGBM. Figure 6b demonstrates that MV2LightGBM identifies the highest number of true positive cases compared to the other models. Additionally, its AUC score of 0.92 highlights its efficiency in distinguishing between positive and negative cases.

MV2LightGBM achieved an accuracy of 91.49%, the highest among the models based on the MobileNetV2 architecture. Although its precision score (86.96%) is slightly lower than that of MV2AdaBoost (MobileNetV2 with AdaBoost) (88.89%), it outperformed other models in terms of the MCC score (0.83). Furthermore, MV2LightGBM attained the highest accuracy among all the models listed in Table 9, Table 10, and Table 11.

86.96% Precision score of MV2LightGBM indicates its ability to identify true positive cases of Mpox, which is higher than MV1LR. Furthermore, its weighted precision of 91.87% shows an overall scenario of how MV2LightGBM is suitable for binary classification, especially for an imbalanced dataset. In addition to that, its 91.87% recall and 95.24% weighted recall highlighted the robustness of this model in detecting true positive cases out of all true positive cases. More specifically, one can see that this model identified 20 Mpox images out of 21 (Figure 6b), which is the highest number of predictions over all other models.

Next, the 90.91% F1 score and the 91.51% weighted F1 score of MV2LightGBM highlighted the strength of this model in minimizing false positive and false negative cases. In other words, it can correctly identify 90.91% of the true positive instances while keeping the false positive and false negative cases low.

Furthermore, MV2LightGBM outperformed all the models in terms of AUC score, demonstrating its superior ability to distinguish between classes. Specifically, MV2LightGBM achieved an AUC of 0.92, which is significantly higher compared to several other models. Its AUC score is 15% higher than XcepSVM, 9% higher than D121LightGBM, 8% higher than MV1LR and IRV2LR. Moreover, its AUC is 4% higher than D169LR and IV3AdaBoost. These results along with its MCC score (0.83) established this model as an efficient robust model for Mpox detection.

### 4.1. Comparison with previous studies

Through our analysis, MV2LightGBM emerged as the best-performing hybrid model, achieving the highest accuracy compared to all other evaluated hybrid models. This section compares this model with the different models previously published in the literature for Mpox detection.

In this paper, we have used the dataset created by [5]. However, MV2LightGBM, which obtained 91.49% accuracy, outperformed the model proposed by [5]. Note that the total augmented images reported in [5] is 2, 562, which is much larger than our total augmented images.

Additionally, they did not conduct any qualitative or quantitative validation tests to ensure their models did not produce any biases, moreover they only computed accuracy, precision, recall and F1 scores. In contrast, we have performed a quantitative validation test on our augmented images and performed a comprehensive analysis by computing various evaluation metrics.

MV2LightGBM also outperformed the model proposed by [58] by a slight margin. Note that [58] utilized the same dataset, including augmented images from [5] and did not provide any evidence of quantitative or qualitative validation.

Furthermore, MV2LightGBM outperformed [62] not only in terms of accuracy but also across other evaluation metrics (see Table 12). [62] trained their models on a different dataset than ours, using a total of 1,754 images, of which 587 are Mpox. However, they did not compute any additional metrics beyond the four we reported in Table 12, despite their dataset being imbalanced.

**Table 12.** Comparison of our best-performing model with previously published results.

|  | Accuracy | Precision | Recall | F1 score |
| --- | --- | --- | --- | --- |
| MV2LightGBM | 91.49 | 86.96 | 95.24 | 90.91 |
| [5] | 82.96 | 87 | 83 | 84 |
| [58] | 91.11 | 90 | 90 | 90 |
| [62] | 87.13 | 85.44 | 85.47 | 85.40 |
| [23] | 87 | 92 | 87 | 90 |

Although MV2LightGBM outperformed the best-performing model, EfficientNetB3, proposed by [23] in terms of accuracy, [23] achieved higher precision than MV2LightGBM. It is important to note that their augmented dataset contains 847 images, of which 160 are Mpox. Moreover, they did not report any validation results to confirm that the augmented images are free from bias. However, MV2LightGBM achieved higher recall and F1 scores compared to [23].

It is important to note that, compared to us, all these prior studies had a larger amount of training data. We explored some DL architectures that contain over 200 layers, as well as architectures suitable for mobile devices, such as MobileNetV1 and MobileNetV2. Moreover, to train models, we did not have to utilize any high-performance computational resources, which highlights the efficiency of our proposed framework in a resource-limited setting.

## 5. Conclusion

A deep learning framework for Mpox detection, integrating pre-trained deep learning architectures and machine learning classifiers, was developed in this paper. This framework was designed as a tool for generating robust models when the amount of data is limited. The input data to the models are skin lesion images, which include Mpox and other skin disease images. To enhance model robustness, data augmentation was applied to the training data. Results from the Kullback-Leibler divergence analysis showed that data augmentation did not significantly alter the data distribution. Using various deep learning architectures, such as DenseNet, Inception, Xception, MobileNet along with nine machine learning classifiers a total of 72 models were trained, followed by a comprehensive analysis. To ensure that the models efficiently analyzed the imbalanced data, various evaluation metrics were computed. The results showed that MV2LightGBM achieved the highest accuracy of 91.49%. Since imbalanced data can lead to erroneous accuracy, MCC scores were given more priority over other metrics in this paper. The MCC results showed that MV2LightGBM achieved the highest score 0.83 over other models, which highlights its reliability in handling imbalanced data. Moreover, the AUC of MV2LightGBM (0.92) further confirms its strong classification capability. These findings demonstrate the potential of the framework for Mpox detection. Further research could explore extending this deep learning framework across diverse datasets in resource-limited settings.

## Data availability

The data we have used to plot Figure 1c is available in Centers for Disease Control and Prevention and Our world data website.

